# Effects of Sudarshan Kriya Yoga (SKY) Breath Meditation on Heart Rate Variability, Sleep and Mental Health in Healthy Individuals : A Pilot and Feasibility Randomized Controlled Trial

**DOI:** 10.1101/2025.01.04.25319992

**Authors:** Vinitha Ganesan, Lauren A Fowler, Sahil Bajaj, Haiyan Qu, Priyam Singh, Snehaa D Krishnan, Keith M McGregor, Timiya S Nolan, Navneet Baidwan, Tapan S Mehta, Ritu Aneja

**Affiliations:** Department of Nutrition Sciences, The University of Alabama at Birmingham, Birmingham, AL, USA; Department of Family and Community Medicine, The University of Alabama at Birmingham, Birmingham, AL, USA; Department of Cancer Systems Imaging, Division of Diagnostic Imaging, The University of Texas MD Anderson Cancer Center, Houston, TX, USA; Department of Health Services Administration, The University of Alabama at Birmingham, Birmingham, AL, USA; Department of Clinical Diagnostics Sciences, The University of Alabama at Birmingham, Birmingham, AL, USA; Division of Preventive Medicine, Department of Medicine, The University of Alabama at Birmingham, Birmingham, AL, USA

**Author notes:** **Correspondence:** Ritu Aneja, PhD; Tapan S Mehta, PhD; The University of Alabama at Birmingham, Birmingham, AL, USA. Data Sharing Statement: De-identified individual participant data and statistical/analytic codes will be shared for secondary analyses upon reasonable request to the corresponding authors.

**Keywords:** Sudarshan Kriya Yoga, SKY, breathwork, meditation, heart rate variability

## Abstract

Effective interventions are needed to address increasing mental health challenges that affect wellbeing. Sudarshan Kriya Yoga (SKY), a breath-based meditation, has previously shown to improve mental health. We conducted a pilot randomized controlled trial (RCT) in healthy individuals, to evaluate the feasibility of SKY intervention with an eight-week followup for studying its immediate and sustained effects on psychophysiological outcomes in real-world settings. 187 healthy individuals (>21 years old) were assessed for eligibility through telephone interviews and 45 consented individuals were randomized (2:1) to either receive three-day SKY intervention with eight-week followup or waitlist control group. The mean age was 40.9 years (SD = 11.3), and 35 (77.8%) were female. Adherence (≥ three-times/week) to 30-minute daily (75.9%) and (≥ six-times/week) 75-minute (89.7%) SKY practice were high. Compared to controls, SKY participants exhibited significantly higher resting heart rate variability (mean difference = 6.47 ([95% CI [0.95, 11.98]) at the end of three-days, and at eight-week followup (mean difference = 5.55 (90% CI [0.46, 10.64]) relative to baseline. At eight-week followup, SKY participants had significant improvements in resting heart rate, overall sleep score, anxiety and social connectedness compared to controls. These results demonstrate feasibility that consistent SKY practice can impact physiology. ClinicalTrials.gov, <u>NCT05523414.</u>

## INTRODUCTION

In 2023, the National Institute of Mental Health (NIMH) reported that 57.8 million adults in the United States live with mental illness, of which anxiety disorders are the most common.^1^ In 2019, 10.8% of US adults had symptoms of anxiety disorder or depressive disorder, and this percentage has increased to 20.5% in 2024.^2^ The rising prevalence of anxiety and depressive disorders underscores the need for effective and accessible interventions to promote mental well-being. Many cultures rely on the calming effects of breathing exercises, the value of which has increasingly been recognized in modern clinical practice.^3^ Recent studies have focused on mind-body practices, including controlled breathing exercises (ie, breathwork), as a potential therapeutic strategy.^4,5^ There have also been studies of the physiology of slow-paced breathing techniques rooted in the yogic tradition.^6^ Neurophysiological studies indicate that high-ventilation breathwork (ie, fast breathing) is associated with extraordinary subjective experience and affects the function of the central and autonomic nervous systems.^7^

Sudarshan Kriya Yoga (SKY), a practice rooted in yogic tradition, includes rhythmic cyclic slow, medium, and fast-paced breathing. Its components include prolonged exhalation, high-ventilation breathwork, and non-sleep deep rest which alleviate stress and enhance overall psychological health and well-being.^8,9^ In a randomized controlled trial (RCT), SKY was found to improve the well-being and social connectedness of university students compared to mindfulness-based stress reduction and emotional intelligence interventions, as determined through subjective self-reported behavioral outcomes.^8^ As a physiological metric, heart rate variability (HRV) can be used as an objective indicator of well-being, providing valuable insights into the relationship between the autonomic nervous system and affective states, including stress.^10,11^ Acute HRV regulation has been studied in SKY practitioners during 75-minute guided group sessions, typically performed in a controlled setting in the presence of a trained instructor.^12–14^ However, the physiological effects of consistent daily SKY home practice in real-world settings remain unknown.

Despite the promising psychophysiological effects of breathwork, the transcriptomic and epigenomic basis for its benefits are largely unknown relative to other forms of meditation.^15^ Small observational studies involving 14 to 47 participants reported reductions in molecular biomarkers of inflammation and stress among SKY practitioners but RCTs have not been undertaken for investigating molecular changes due to SKY breath meditation.^16–20^ Therefore, robust and well-designed RCT are needed to understand the molecular basis for breathwork-associated improvements in psychophysiological well-being.^21^ In this pilot study, we report the primary and secondary outcomes of the effects of SKY breath meditation on psychophysiological well-being in a real-world setting. We also discuss the feasibility of gathering real-world data along with blood collection to help inform the development of future clinical trials investigating molecular mechanisms underlying these effects.

## METHODS

### Participants

Participants were recruited primarily from San Diego, CA through the Art of Living website, email newsletters, social media, and word of mouth. Study recruitment fliers were posted via the online platforms, and individuals were invited to apply by filling out a brief interest form. Those who completed the form were contacted by a research team member and screened for eligibility via telephone.

A detailed description of inclusion and exclusion criteria is provided in the supplement. Briefly, inclusion criteria were >21yrs of age, ≥ 5^th^ grade competency in English, and agreement to complete ≥ 4 weekly SKY practice sessions during followup. Exclusion criteria included diagnosis of major medical or psychiatric condition; major surgery within last 12-weeks; current tobacco/heavy alcohol use; recent or current substance abuse; taking lithium-based medication or hormone-replacement therapy; pregnant/breastfeeding; current participation in similar studies; regular practice of meditation or breath-based technique (≥ 3-times/week). The inclusion and exclusion criteria were intentionally restrictive, keeping in mind the multi-omic analysis for mechanistic exploration, as pre-existing conditions, medications, alcohol consumption, substance use, and smoking may affect molecular signatures.

Informed consent was obtained from all participants prior to initiation of the study. The trial protocol was registered at clinicaltrials.gov (<u>NCT05523414</u>, registered 31/08/202) and approved by the UAB Institutional Review Board. All methods were performed in accordance with the guidelines and regulations from the UAB Insitutional Review Board. This report follows the Consolidated Standards of Reporting Trials (CONSORT) guidelines for RCT.^22^

### Study Design and Randomization

This pilot study was a parallel, two-group RCT, in which participants were assigned to either the SKY intervention or a waitlist control arm over an eight-week period. Participants randomized to the control group were provided the opportunity to participate in SKY after completion of the study. Participants were randomized to the intervention and control arms at a ratio of 2:1, respectively. Our choice of sample size was based on having a minimum of 12 participants per arm, as previously recommended for external pilot studies,^23^ whereas the allocation ratio of 2:1 was selected to prioritize piloting the intervention arm.

### SKY Intervention

SKY breath meditation consists of three different breathing techniques: (1) three-stage victory breath, (2) bellows breath, and (3) SKY breath, which is a cyclical breathing protocol guided by an audio recording consisting of slow, medium, and fast breath cycle rates.^24^ The SKY intervention was administered using a group format in a meditation center by two SKY instructors from the Art of Living Foundation (a registered non-profit organization). Group sessions involved approximately 21 contact hours. Prior to followup, participants attended a three-day intensive group workshop ( three-hours/day) called the Art of Living Part 1 Course. Participants learned both a 75-minute guided SKY practice (‘long SKY’) to be performed once-weekly in a group class and 30-minute home SKY practice (‘home SKY’) to be performed daily. Participants attended eight in-person long SKY sessions during followup, with a guided 60-minute session offered via Zoom for those who could not attend in-person. To support daily SKY practice at home, a smartphone application called “Art of Living Journey” was offered free of charge.

### Data Collection

Detailed descriptions of data collection procedures and monetary incentives are provided in eMethods, Supplement 1. At baseline, all participants were issued a Garmin Vivosmart 5 smartwatch and instructed to install the Labfront and Garmin Connect smartphone applications (required for collection of SKY adherence logs and real-world physiological data). Participants were directed to wear the watch for the duration of the study. Behavioral surveys were administered via REDCap at baseline and eight-weeks post-intervention. Demographic data (age, sex, race, ethnicity, body weight, and height) were also collected as part of the REDCap survey. To perform multi-omic analysis, blood was collected at baseline, at the end of three-day SKY workshop, and at the end of eight-week followup from both groups.

### Study outcomes

Primary outcomes were feasibility metrics and measures for HRV and respiratory rate. Secondary outcomes included self-reported behavioral measures and additional physiological measures. Other outcomes included blood biochemistry, epigenetics, transcriptomics and proteomics investigation of blood samples collected from this study, analysis of which are underway. Full descriptions of each outcome are provided in eMethods, Supplement 1.

#### Feasibility Metrics

Feasibility was assessed by examining recruitment and refusal rates among those screened for eligibility, and rates of retention, wearable device compliance, and intervention adherence in randomized participants.

#### Physiological Outcomes

The Garmin watch was used to collect data for daily resting heart rate, respiration rate, HRV (resting and awake), and overall sleep scores at baseline and during the eight-week study period.

#### Behavioral Outcomes

Self-reported measures related to mental health, psychological thriving, and personality were evaluated in accordance with a previous study that investigated effects of SKY in students in a university setting (eTable 1 in Supplement 1).^8^

### Statistical Analysis

Analyses for feasibility and physiological outcomes were conducted in R 4.3.3 (R Core Team, 2024)^25^ with the following packages: tidyverse v2.0.0,^26^ DescTools v0.99.54,^27^ gtsummary v1.7.2,^28^ ggfortify v0.4.17,^29^ car v3.1-2,^30^ ggeffects v1.5.2, and effectsize v0.8.7.^31^ Behavioral outcomes were analyzed using IBM SPSS Version 29 (IBM Corp, Armonk, NY).^32^ Continuous variables were reported as mean (SD) or median (minimum/maximum values), whereas categorical variables were reported as frequency (%). Feasibility metrics and baseline characteristics were evaluated using descriptive analyses. All other analyses are detailed in eMethods, Supplement 1. All tests reported findings at significance levels of 0.05, 0.10 and 0.20 (ie at 95%, 90% and 80% confidence intervals [CIs]) in line with the statistical interpretation of pilot studies ^33^.

## RESULTS

### Study Participants

Counts for participant enrollment, allocation, intervention, and followup are shown in Figure 1. Of the 187 individuals assessed for eligibility, 68 (36.4%) were eligible, of whom 45 (24.1%) consented to participate in the study and were randomized. The baseline characteristics of the enrolled participants are presented in Table 1. The mean (SD) age of the overall sample was 40.9 (11.3) years, and 35 (77%) participants were female.

**Figure 1.**
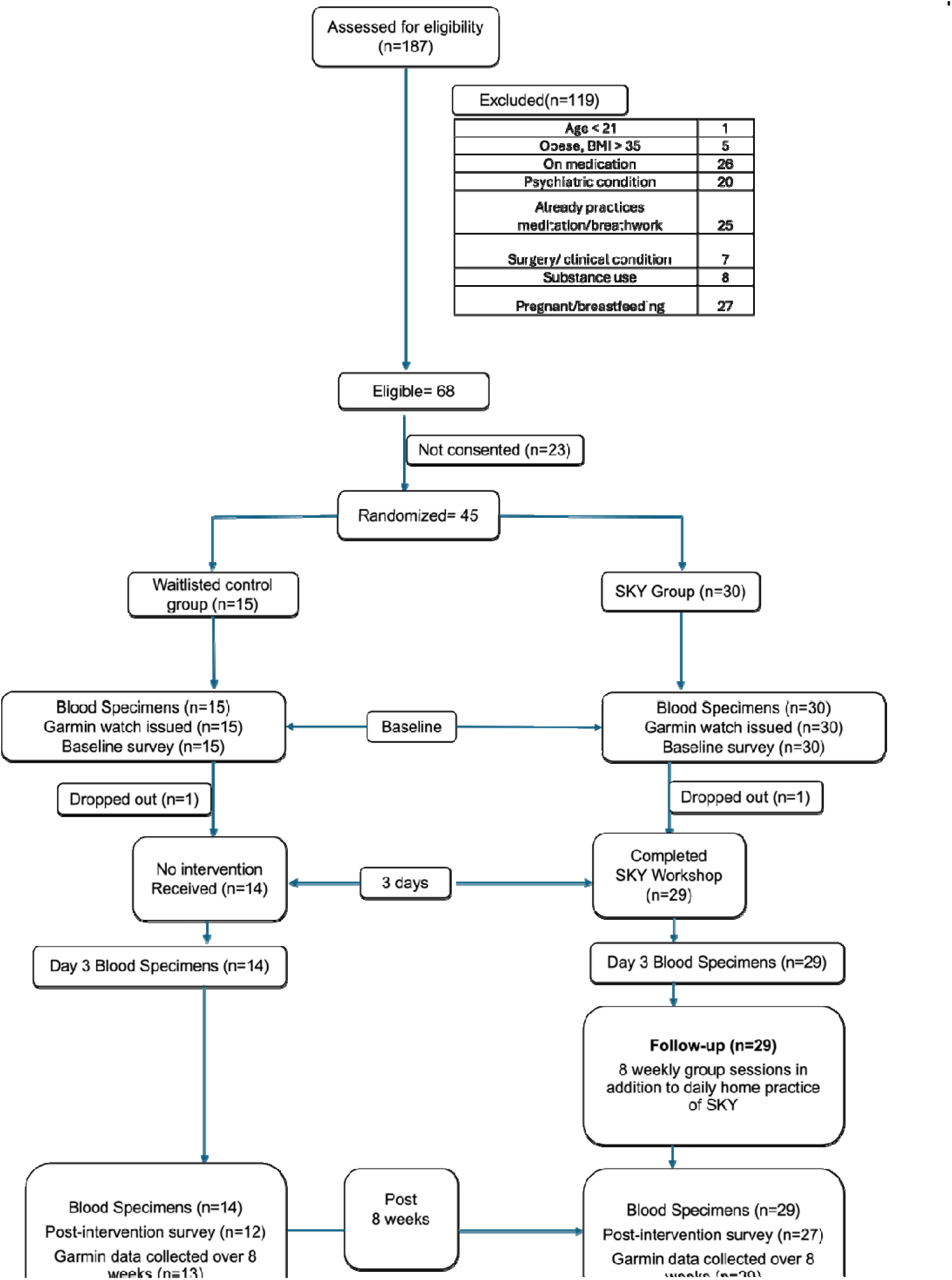
Consolidated Standards of Reporting Trials Flow Diagram of This Parallel Randomized Trial of Individuals Assigned to the SKY Intervention or Waitlisted Control Groups

**Table 1.**
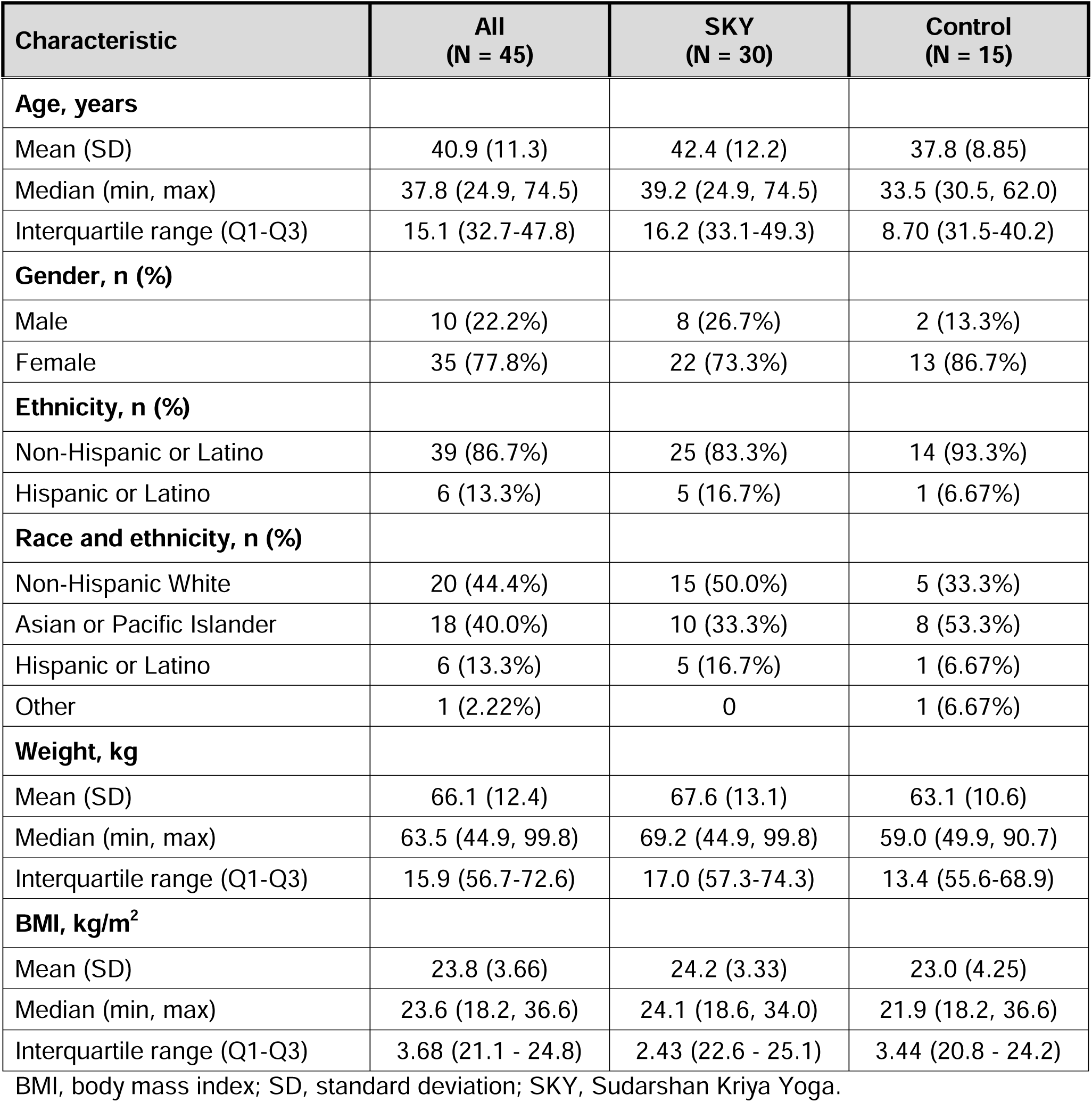
Baseline Demographic and Clinical Characteristics in all Participants and by Treatment Group. Data are expressed as *N* participants (%) unless indicated otherwise. ^a^Descriptive summaries are computed from the average number of daily practice sessions performed per week over the followup period for each participant.SD, standard deviation; SKY, Sudarshan Kriya Yoga.

| Characteristic | All<br>(N = 45) | SKY<br>(N = 30) | Control<br>(N = 15) |
| --- | --- | --- | --- |
| **Age, years** |  |  |  |
| Mean (SD) | 40.9 (11.3) | 42.4 (12.2) | 37.8 (8.85) |
| Median (min, max) | 37.8 (24.9, 74.5) | 39.2 (24.9, 74.5) | 33.5 (30.5, 62.0) |
| Interquartile range (Q1-Q3) | 15.1 (32.7-47.8) | 16.2 (33.1-49.3) | 8.70 (31.5-40.2) |
| **Gender, n (%)** |  |  |  |
| Male | 10 (22.2%) | 8 (26.7%) | 2 (13.3%) |
| Female | 35 (77.8%) | 22 (73.3%) | 13 (86.7%) |
| **Ethnicity, n (%)** |  |  |  |
| Non-Hispanic or Latino | 39 (86.7%) | 25 (83.3%) | 14 (93.3%) |
| Hispanic or Latino | 6 (13.3%) | 5 (16.7%) | 1 (6.67%) |
| **Race and ethnicity, n (%)** |  |  |  |
| Non-Hispanic White | 20 (44.4%) | 15 (50.0%) | 5 (33.3%) |
| Asian or Pacific Islander | 18 (40.0%) | 10 (33.3%) | 8 (53.3%) |
| Hispanic or Latino | 6 (13.3%) | 5 (16.7%) | 1 (6.67%) |
| Other | 1 (2.22%) | 0 | 1 (6.67%) |
| **Weight, kg** |  |  |  |
| Mean (SD) | 66.1 (12.4) | 67.6 (13.1) | 63.1 (10.6) |
| Median (min, max) | 63.5 (44.9, 99.8) | 69.2 (44.9, 99.8) | 59.0 (49.9, 90.7) |
| Interquartile range (Q1-Q3) | 15.9 (56.7-72.6) | 17.0 (57.3-74.3) | 13.4 (55.6-68.9) |
| **BMI, kg/m<sup>2</sup>** |  |  |  |
| Mean (SD) | 23.8 (3.66) | 24.2 (3.33) | 23.0 (4.25) |
| Median (min, max) | 23.6 (18.2, 36.6) | 24.1 (18.6, 34.0) | 21.9 (18.2, 36.6) |
| Interquartile range (Q1-Q3) | 3.68 (21.1 - 24.8) | 2.43 (22.6 - 25.1) | 3.44 (20.8 - 24.2) |

BMI, body mass index; SD, standard deviation; SKY, Sudarshan Kriya Yoga.

### Feasibility Metrics

#### Recruitment and Refusal rates

One-third of the eligible individuals (23/68) either declined to participate because of scheduling conflicts or did not return the signed consent forms, the reasoning for refusal could not be documented clearly as there were multiple modes of communication during the recruitment process via phone calls, text messages and email.

#### Study Completion Rates

One participant in the SKY group (1/30; 3.33%) dropped out during the workshop, and one participant in the control group (1/15; 6.67%) dropped out between randomization and the start of the followup. Overall, 43 participants (95.6%) completed the pre-intervention behavioral questionnaire, and 38 (84.4%) completed both the pre- and post-intervention questionnaires. Within the SKY group, 3 (20.0%) did not complete the post-intervention questionnaire, and 1 (3.33%) did not complete either questionnaire. In the control group, 3 (20.0%) did not complete the post-intervention questionnaire, and 1 (6.67%) did not complete either questionnaire. Blood samples were collected from 43 (95.6%) participants who completed the study and multi-omic analysis is underway. Hence further investigation using a multi-omic approach is feasible. Overall, 36 (80.0%) participants received all monetary installments (SKY: 24/30 [80.0%]; control: 12/15 [80.0%]).

#### Garmin watch compliance

The Garmin watch was worn by 42/45 participants (93.3%) randomized at baseline (SKY, 29 [96.7%]; control, 13 [86.7%]). Results reported for wearable device compliance pertain to these 42 participants. Wearable compliance during sleep was notably lower relative to compliance during awake periods (Supplement 2, eTable 7).

#### Adherence rates

Adherence to daily SKY practice during followup is detailed in Table 2 and eTable 2 (Supplement 2). SKY participants performed the daily home SKY practice an average of 4.27 (SD = 0.94) days per week during followup. Adherence was highest for weeks 1-2 (100% of all participants) and decreased to 93.1% for weeks 5–8. For the once-weekly guided long SKY sessions (Table 2), SKY participants attended an average of 6.52 (SD = 1.06) total sessions over the followup period. Overall, 26 (89.7%) participants participated in at least six sessions and 7 of 29 (24.1%) attended all eight sessions. Adherence to weekly long SKY practice with the group stayed consistent for the weeks 1-4 (75%), declined slightly in weeks 5 - 6 (68%), followed by an increase to 100% in week 7 and 96% in week 8.

**Table 2.**
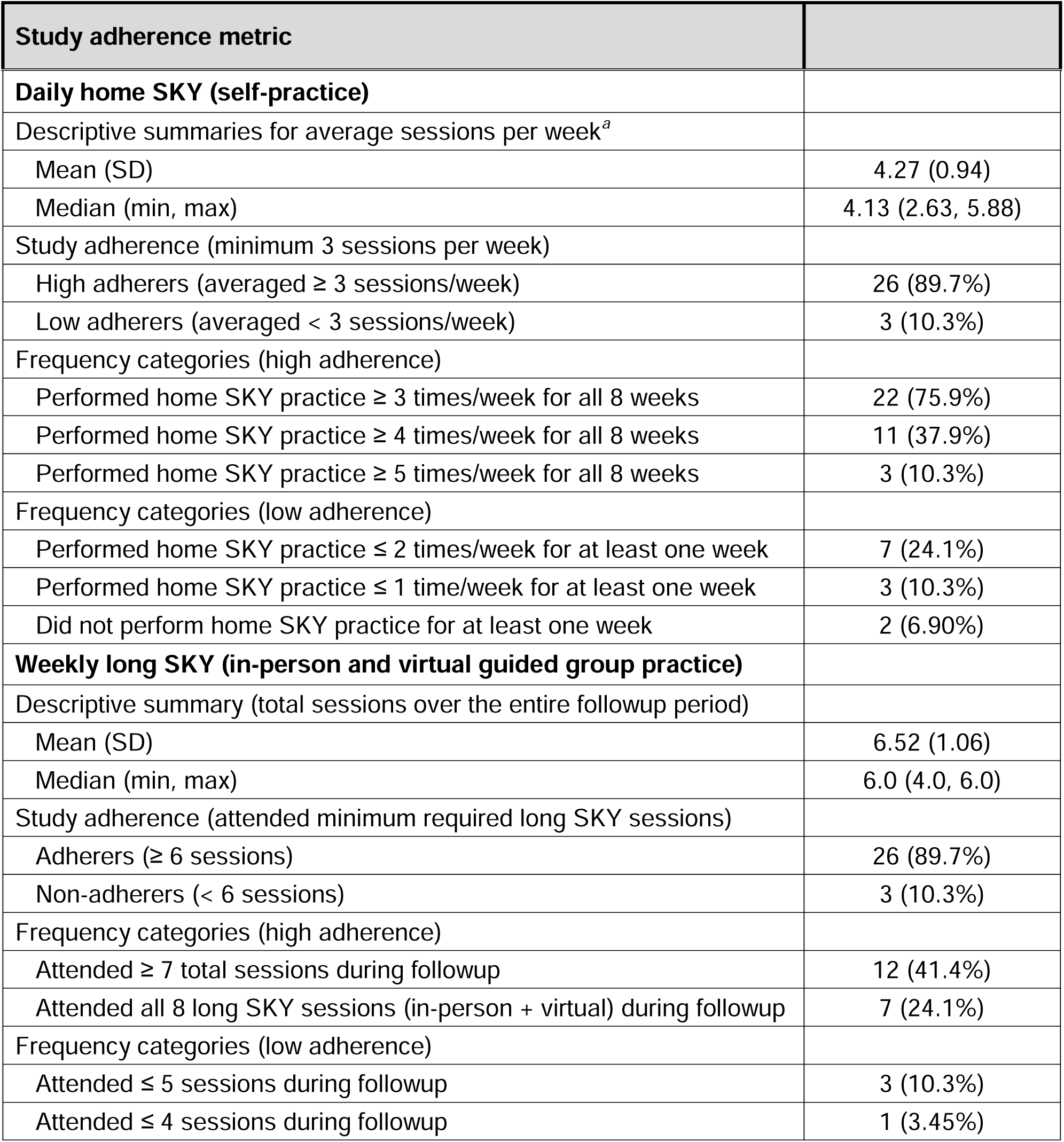
Adherence to Daily Home SKY (30-minute) Practice and Weekly SKY (60-75-minute) Group Practice in the Intervention Group (*N* = 29) Over Eight-week Followup.

### Physiological Outcomes

Between-group differences in Garmin measurements are presented in **Table 3**. On day-3 of the SKY workshop, SKY participants had a lower resting HR (MD = −2.33 [95% CI: −4.76,0.10; 90% CI: −4.37, −0.29]) and higher resting HRV (MD = 6.47 [95% CI: 0.95, 11.98]) compared to baseline. A medium effect of treatment (g = 0.64) was identified for resting HR, whereas a large effect was observed for resting HRV (g = −0.91) immediately following the three-day SKY workshop. At the eight-week followup, there were no notable effects on resting HR; SKY participants had sustained a higher resting HRV (MD = 5.55 [95% CI: −0.51,11.6; 90% CI: 0.46, 11.98]) compared to baseline. A medium effect of treatment (g = −0.81) was identified for resting HRV at the eight-week followup. While there were no immediately apparent effects on overall sleep score at the end of the three-day workshop, a medium effect of treatment (g=-0.82) was identified for overall sleep score (MD = 8.88 [95%CI: 0-0.64,18.4; 90%CI: 0.89, 16.86]) at the eight-week followup compared to baseline.

**Table 3.**
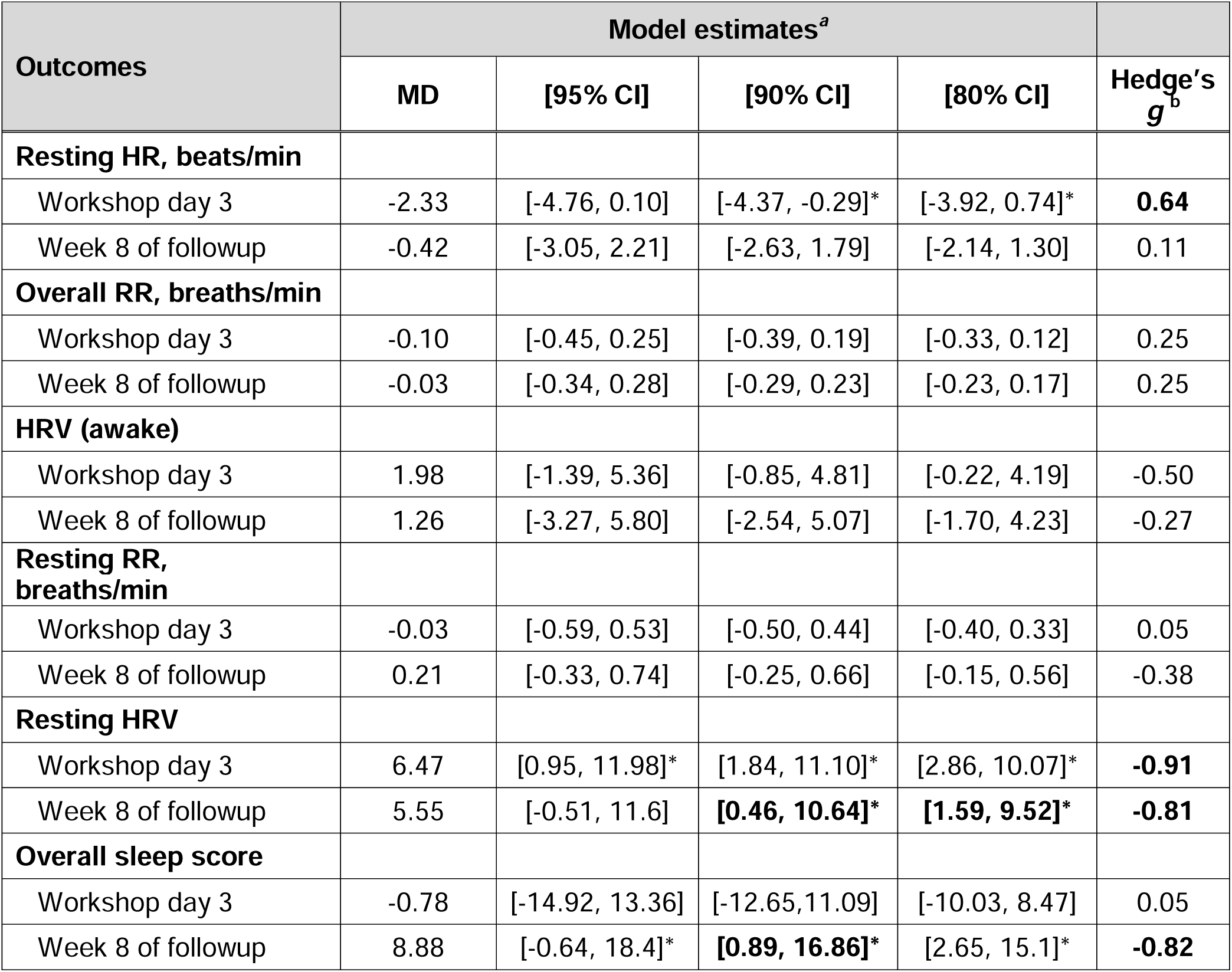
Between-group Estimates for Physiological Outcomes on Day-3 of the SKY Workshop and Week 8 of Followup. *^a^*Derived from general linear regression models and adjusted for baseline values. Estimates [CI] are for the SKY group and reflect the mean difference from the control group (reference). *^b^*Interpretation of Hedge’s *g* values: very small (*g* <0.2); small (0.2≤ *g <*0.5); medium (0.5 ≤ *g* <0.8); large (*g* ≥ 0.8). Starred [CI] = Interval does not include 0. CI, confidence interval; HR, heart rate; HRV, heart rate variability; LSM, least square means; RR, respiration rate; MD, Mean Difference;

| Outcomes | Model estimates<sup>a</sup> | | | | Hedge's $g^b$ |
| --- | --- | --- | --- | --- | --- |
|  | MD | [95% CI] | [90% CI] | [80% CI] |  |
| **Resting HR, beats/min** |  |  |  |  |  |
| Workshop day 3 | -2.33 | [-4.76, 0.10] | [-4.37, -0.29]* | [-3.92, 0.74]* | **0.64** |
| Week 8 of followup | -0.42 | [-3.05, 2.21] | [-2.63, 1.79] | [-2.14, 1.30] | 0.11 |
| **Overall RR, breaths/min** |  |  |  |  |  |
| Workshop day 3 | -0.10 | [-0.45, 0.25] | [-0.39, 0.19] | [-0.33, 0.12] | 0.25 |
| Week 8 of followup | -0.03 | [-0.34, 0.28] | [-0.29, 0.23] | [-0.23, 0.17] | 0.25 |
| **HRV (awake)** |  |  |  |  |  |
| Workshop day 3 | 1.98 | [-1.39, 5.36] | [-0.85, 4.81] | [-0.22, 4.19] | -0.50 |
| Week 8 of followup | 1.26 | [-3.27, 5.80] | [-2.54, 5.07] | [-1.70, 4.23] | -0.27 |
| **Resting RR, breaths/min** |  |  |  |  |  |
| Workshop day 3 | -0.03 | [-0.59, 0.53] | [-0.50, 0.44] | [-0.40, 0.33] | 0.05 |
| Week 8 of followup | 0.21 | [-0.33, 0.74] | [-0.25, 0.66] | [-0.15, 0.56] | -0.38 |
| **Resting HRV** |  |  |  |  |  |
| Workshop day 3 | 6.47 | [0.95, 11.98]* | [1.84, 11.10]* | [2.86, 10.07]* | **-0.91** |
| Week 8 of followup | 5.55 | [-0.51, 11.6] | **[0.46, 10.64]\*** | **[1.59, 9.52]\*** | **-0.81** |
| **Overall sleep score** |  |  |  |  |  |
| Workshop day 3 | -0.78 | [-14.92, 13.36] | [-12.65, 11.09] | [-10.03, 8.47] | 0.05 |
| Week 8 of followup | 8.88 | [-0.64, 18.4]* | **[0.89, 16.86]\*** | [2.65, 15.1]* | **-0.82** |

No evidence of a treatment effect was observed for other physiological outcomes (overall respiratory rate, resting respiratory rate, HRV during awake periods) on day-3 of workshop or eight-week followup. Descriptive summaries of Garmin outcomes for baseline, three-days of SKY workshop, and each week of followup are presented in eTables 4 and 5 in Supplement 2. Marginal estimates from the generalized linear models are provided in eTable 6 in Supplement 2.

### Behavioral Outcomes

Between-group differences in behavioral survey responses at eight-week followup are presented in Table 4. Marginal estimates from the generalized linear models are provided in eTable 3 in Supplement 2.

**Table 4.** Between-group Comparisons of Behavioral Outcomes at Week 8 (SKY, N=26 and Control, N=13). *^a^*Derived from general linear regression models and adjusted for baseline scores. Estimates [CI] are for the SKY group and reflect differences from the control group (reference). *^b^*Adj. *P*-values adjusted for multiple comparisons using the Benjamini and Hochberg method. *P* <0.10 was considered significant. *^c^*Interpretation of Hedge’s *g* values: very small (*g* <0.2); small (0.2 *g <*0.5); medium (0.5≤ *g* < 0.8); large (*g* ≥ 0.8). *[CI] = Interval does not include 0. BFI-44, Big Five Inventory; Brief-COPE, Coping Orientation to Problems Experienced Inventory; CI, confidence interval; MASQ D30, Mood and Anxiety Symptom Questionnaire; PANAS, Positive and Negative Affect Schedule; RYFF-18, Psychological Well-Being Scale; MD, Mean Difference;

| Outcome | Model estimates<sup>a</sup> | | | | | | Hedge's $g^c$ |
| --- | --- | --- | --- | --- | --- | --- | --- |
|  | MD | [95% CI] | [90% CI] | [80% CI] | *P* value | Adj<sup>b</sup> *P* |  |
| Burnout | -0.32 | [-0.74, 0.10] | [-0.68, 0.03] | [-0.60, -0.05]* | 0.141 | 0.296 | 0.52 |
| Perceived stress | -3.53 | [-6.19,<br>-0.86]* | [-5.77,<br>-1.29]* | [-5.27,<br>-1.78]* | **0.014** | **0.083** | 0.90 |
| **Mood and Anxiety Symptoms (MASQ D30)** |  |  |  |  |  |  |  |
| Overall score | -10.5 | [-16.5,<br>-4.50]* | [-15.5,<br>-5.46]* | [-14.4,<br>-6.56]* | **0.002** | **0.025** | 1.18 |
| General distress | -3.24 | [-6.59,<br>0.12] | [-6.05,<br>-0.42]* | [-5.43,<br>-1.05]* | **0.066** | 0.199 | 0.65 |
| Anxious arousal | -3.50 | [-5.28,<br>-1.72]* | [-5.00,<br>-2.01]* | [-4.67,<br>-2.34]* | **<0.001** | **0.015** | 1.37 |
| Anhedonic depression | -3.11 | [-6.75,<br>0.53] | [-6.17,<br>-0.06]* | [-5.49,<br>-0.73]* | 0.102 | 0.260 | 0.58 |
| **Positive and Negative Emotions (PANAS)** |  |  |  |  |  |  |  |
| Positive affect | 3.95 | [0.85,<br>7.05]* | [1.35,<br>6.55]* | [1.92,<br>5.98]* | **0.017** | **0.083** | -0.86 |
| Negative affect | -1.99 | [-5.21,<br>1.22] | [-4.69,<br>0.71] | [-4.10,<br>0.11] | 0.232 | 0.365 | 0.42 |
| **Psychological Well-Being (RYFF-18)** |  |  |  |  |  |  |  |
| Overall score | 2.16 | [-3.35,<br>7.67] | [-2.46,<br>6.78] | [-1.44,<br>5.76] | 0.447 | 0.614 | -0.27 |
| Autonomy | 0.51 | [-1.18,<br>2.19] | [-0.91,<br>1.92] | [-0.60,<br>1.61] | 0.560 | 0.684 | -0.21 |
| Environmental mastery | 1.27 | [-0.51,<br>3.05] | [-0.22,<br>2.76] | [0.11,<br>2.43]* | 0.171 | 0.296 | -0.48 |
| Personal growth | 0.05 | [-1.02,<br>1.11] | [-0.85,<br>0.94] | [-0.65,<br>0.74] | 0.934 | 0.963 | -0.03 |
| Positive relations with others | -0.02 | [-1.37,<br>1.34] | [-1.16,<br>1.12] | [-0.90,<br>0.87] | 0.978 | 0.978 | 0.000 |
| Purpose in life | 0.12 | [-1.37,<br>1.61] | [-1.13,<br>1.37] | [-0.85,<br>1.09] | 0.876 | 0.932 | -0.05 |
| Self-acceptance | 0.20 | [-1.28,<br>1.67] | [-1.04,<br>1.43] | [-0.77,<br>1.16] | 0.794 | 0.885 | -0.09 |
| Eudaimonic well-being | 1.15 | [0.23,<br>2.06]* | [0.38,<br>1.91]* | [0.55,<br>1.74]* | **0.019** | **0.083** | -0.90 |
| Life satisfaction | 1.38 | [-0.44,<br>3.20] | [-0.15,<br>2.91] | [0.19,<br>2.57]* | 0.145 | 0.296 | -0.53 |
| Gratitude | -0.30 | [-1.98,<br>1.38] | [-1.71,<br>1.11] | [-1.4,<br>0.80] | 0.730 | 0.860 | 0.12 |
| Optimism | 2.06 | [0.11,<br>4.02]* | [0.42,<br>3.70]* | [0.79,<br>3.34]* | **0.046** | 0.151 | -0.72 |
| Self-esteem | 0.44 | [-0.16,<br>1.04] | [-0.06,<br>0.94] | [0.05,<br>0.83]* | 0.159 | 0.296 | -0.53 |
| Self-compassion | 6.01 | [0.85,<br>11.16]<br>* | [1.68,<br>10.33]<br>* | [2.63,<br>9.38]* | **0.028** | 0.104 | -0.83 |
| Social connectedness | 7.47 | [2.30,<br>12.64]<br>* | [3.13,<br>11.81]<br>* | [4.09,<br>10.85]<br>* | **0.008** | **0.083** | -0.98 |
| Mindfulness | 3.92 | [0.89,<br>6.95]* | [1.38,<br>6.46]* | [1.94,<br>5.90]* | **0.016** | **0.083** | -0.89 |

| **Coping styles (Brief-COPE)** |  |  |  |  |  |  |  |
| --- | --- | --- | --- | --- | --- | --- | --- |
| Avoidant coping | -0.82 | [-1.95, 0.32] | [-1.77, 0.14] | [-1.56, -0.07]* | 0.169 | 0.296 | 0.48 |
| Problem-focused | 1.53 | [-0.38, 3.44] | [-0.07, 3.13] | [0.28, 2.78]* | 0.126 | 0.296 | -0.55 |
| Emotion-focused | 0.77 | [-1.63, 3.16] | [-1.25, 2.78] | [-0.80, 2.33] | 0.536 | 0.680 | -0.22 |
| **Personality Factors (BFI-44)** |  |  |  |  |  |  |  |
| Extraversion | 0.22 | [0.04, 0.40]* | [0.07, 0.37]* | [0.10, 0.34]* | **0.015** | **0.083** | -0.84 |
| Agreeableness | 0.16 | [-0.08, 0.40] | [-0.04, 0.36] | [0.01, 0.32]* | 0.194 | 0.320 | -0.46 |
| Conscientiousness | 0.10 | [-0.15, 0.34] | [-0.11, 0.30] | [-0.06, 0.26] | 0.436 | 0.614 | -0.27 |
| Neuroticism | -0.37 | [-0.79, 0.05] | [-0.72, -0.02]* | [-0.64, -0.09]* | **0.093** | 0.156 | 0.60 |
| Openness | 0.09 | [-0.17, 0.34] | [-0.13, 0.30] | [-0.08, 0.25] | 0.511 | 0.675 | -0.23 |
| **Emotion Mindset** |  |  |  |  |  |  |  |
| Emotional intelligence | -0.69 | [-2.43, 1.06] | [-2.15, 0.78] | [-1.83, 0.45] | 0.446 | 0.614 | 0.27 |
| Beliefs/opinions about emotion | -0.38 | [-3.39, 2.63] | [-2.91, 2.14] | [-2.35, 1.59] | 0.804 | 0.885 | 0.09 |

#### Mood, stress, depression and anxiety

Evidence for a large treatment effect were observed in SKY group for perceived stress (MD= −3.53 [95% CI: −6.19, −0.86]; g = 0.90), overall mood and anxiety symptoms score (MASQ D30) (MD = −10.5 [95% CI: −16.5, −4.50], g =1.18), and anxious arousal (MD = −3.50 [95% CI, −5.28, −1.72]; g = 1.37). Evidence of a medium effect were observed in SKY group for general distress (MD = −3.24 [90% CI, −6.05, −0.42], g = 0.65) and anhedonic depression (MD = −3.11[90%CI, −6.17, −0.06], g = 0.58). No evidence of an effect of treatment was observed for burnout measure.

#### Psychological thriving

Post-intervention scores for six measures related to psychological thriving were higher in SKY participants. Evidence for a large effect of treatment was identified for positive affect (MD = 3.95 [95% CI: 0.85, 7.05]; g = 0.86), eudaimonic well-being (MD = 1.15 [95% CI: 0.23, 2.06]; g = 0.90), self-compassion (MD = 6.01 [95% CI: 0.85, 11.86]; g = 0.83), social connectedness (MD = 7.47 [95% CI: 2.30,12.64]; *g* = 0.98), and mindfulness (MD = 3.92 [95% CI: 0.89,6.95]; g = 0.89). A medium effect was observed for optimism (MD = 2.06 [95% CI: 0.11,4.02]; g = 0.72). No evidence of an effect of treatment was observed for measures of negative affectivity, psychological well-being (overall score), gratitude, self-esteem, coping style, emotional mindset.

#### Personality domains

For the personality domains, SKY participants reported higher levels of extraversion (MD= 0.22 [95% CI: 0.04, 0.40]; with a large effect of treatment (g = 0.84) and lower levels of neuroticism (MD = −0.37 [90% CI: −0.72, −0.02]), with a medium effect of treatment (d = 0.60). No evidence of an effect of treatment was observed for any other personality factors.

## DISCUSSION

SKY breath meditation has long been used to support mental health and studied in countries such as India, Norway and Turkey.^16–20^ Although previous studies have shown that SKY can improve subjective measures of mental health and well-being, this is the first study to show the ability of SKY breath meditation to improve resting HRV and sleep in real-world settings with consistent practice. We also found high levels of compliance to both the wearable device and the SKY practice among study participants. These findings suggest a strong commitment to the study protocol, which is critical for the reliability and validity of the research findings.

In this study, physiological measures were continuously collected at two-minute intervals over an eight-week period using Garmin smartwatches, resulting in a high volume of data that enabled analyses of the effects of SKY in both controlled and real-world settings. To examine the effectiveness of SKY, we compared the results at the end of the three-day workshop (indicating the effects in a controlled group setting) with the results at the end of the eight-week followup (indicating the effects in a real-world setting) between intervention and control groups.

HRV measures the relative contribution of the branches of the autonomic nervous system (ie, parasympathetic [PNS] and sympathetic) and is useful as an index of physiological stress. Higher resting HRV is associated with increased engagement of the PNS and may indicate lower chronic stress.^11,34^ Prior studies have shown that rhythmic breathing influences brain connectivity (for instance, synchronizing activity in brain regions involved in emotion and memory processing) and promoting emotion regulation by modulating activity in the limbic system.^35^ Chronic stress alters functional activation and connectivity in interoceptive brain regions, including the insula and amygdala, which are involved in emotional processing and anxiety-related behaviors.^36,37^ Slower breathing enhances brain connectivity between interoceptive and frontal brain networks i.e., the networks that are responsible for executive control of the body state.^38^ SKY breathing likely promotes both autonomic and cognitive regulation. Our finding of increased resting HRV at acute (three-day) and followup (eight-week) assessments is notable. Interestingly, the improvement in resting HRV on a weekly-basis correlated with adherence rates of long SKY practice. Previous studies have reported increases in HRV measured through electrocardiogram (ECG) and optical pulse readings during waking hours.^39,40^ Acute effects of SKY practice on HRV have been documented in controlled settings through ECG measurements.^12–14,41–43^ However, little is known about the relationship between HRV during sleep and vagal tone, particularly concerning the effects of somatic control of breathing. A systematic review of breathwork studies using slow-paced breathing techniques^38^, has identified only one study^44^ that measured changes in resting HRV during sleep, documenting significant alterations in resting HRV only if the practice immediately follows sleep. The sustained increase in resting HRV through week 8 of our study, regardless of time of SKY practice indicates a shift toward PNS and autonomic cardiac tone. A previous study^45^ showed that a mindfulness meditation program involving breath awareness increased HRV during sleep for over 30 sessions. However, we believe that our study is the first to show sustained HRV improvement over two months.

After the eight-week followup, the overall sleep score in the SKY group showed notable improvement. This improvement in sleep quality over time indicates that the beneficial effects of the SKY intervention on sleep become more pronounced with sustained practice, as no significant improvement was observed immediately following the three-day intervention. This delayed benefit suggests that a cumulative effect may be necessary to improve sleep health. The potential mechanisms underlying this improvement in sleep may be related to the ability of SKY to modulate the autonomic nervous function. Previous findings suggest that SKY influences both the PNS and sympathetic branches, promoting relaxation and reducing stress, which are key factors in improving sleep quality.^24,46^ SKY may help balance the hypothalamic-pituitary-adrenal axis, reducing cortisol levels, and consequently, physiological arousal.^24^ This can alleviate hyperarousal, which is often associated with sleep disturbances such as insomnia. Additionally, SKY’s rhythmic breathing techniques enhance vagal tone and stimulate the PNS, which supports restorative sleep and overall physiological relaxation.^46^ Although previous studies involving acute and long-term exercise interventions have shown improvements in HRV during sleep, the autonomic benefits of physical activity involve numerous systemic changes, including improved respiratory, circulatory, neurological, and endocrine function.^47–49^ We hypothesize that there may be systemic changes in neurological and endocrine function associated with SKY breathing that can be investigated in our exploratory multi-omic analysis and future confirmatory studies.

A 2020 study showed that the SKY breath meditation improved depression, stress, mental health, mindfulness, positive affect, and social connectedness compared to mindfulness-based stress reduction and emotional intelligence interventions in university students.^8^ Our results align with the findings of Seppala et al.^8^ and support additional behavioral improvements, including general distress, depression, anxiety, anxious arousal, eudaimonic well-being, extraversion, and neuroticism. Brief structured respiration practices, through various breathwork modalities, have been previously linked to improvements in positive affectivity, and improvements in positive affectivity were associated with higher adherence to breathwork practice.^50^ We also observed improvements in positive affectivity in our study, supporting the hypothesis that intentional control over breath with specific breathing patterns provides greater mood improvements than passive attention to breath (eg, mindfulness meditation). A recent meta-analysis of RCTs on breathwork interventions showed that breathwork was associated with reduced post-intervention self-reported stress levels.^5^ It may be worthwhile to explore the effects of SKY breath meditation on outcomes that showed large effect sizes (eg, perceived stress, depression, anxiety, social connectedness) in larger studies. These results are critical and timely, considering the rising rates of loneliness and isolation in the United States.^51^ Further qualitative research is needed to identify the aspects of the SKY intervention that improve social connectedness and determine whether improvements in social connectedness precede benefits to health outcomes and well-being.

## Limitations

Although the findings of this study are promising, there are several limitations. As SKY is a group intervention, the availability of individuals to participate in the workshop at the same time may be a barrier to participation. In a larger trial, participant recruitment and intervention delivery would be feasible if multiple scheduling options are provided over several weeks or months. This study was proposed as feasibility and pilot study; hence, the study would be underpowered to detect small to moderate effects and may be susceptible to winner’s curse in terms of large effects detected.^52^ Moreover, the limited recruitment channels and sampling procedures hindered the recruitment of a diverse population, thereby reducing the generalizability of the results. For example, Black Americans were not recruited in the pilot study. To ensure that future studies are more inclusive and representative, the study team should be more diverse, and engage community stakeholders in designing and conducting the research and sharing the data with the community for future positive impacts.^53,54^ Factors such as economic instability, neighborhood disadvantage, and food insecurity can also affect mental health and well-being; hence, demographic details such as education, income levels, and economic stability should be collected in future studies. In addition, the data collected during sleep cycles may be limited because many participants exhibited lower compliance with wearing the Garmin wristwatches during sleep. Consequently, findings related to sleep patterns and quality should be interpreted with caution. Detailed sleep studies are needed to gain a more comprehensive and accurate understanding of the physiological mechanisms involved in the impact of the SKY intervention on sleep health.

## CONCLUSIONS

The high completion rates strongly support the feasibility of the eight-week SKY study. High compliance with wearable device use and good adherence to SKY (both long and home practice) were observed. Improvements in resting HRV and overall sleep scores support the positive physiological effects of the SKY intervention and the significant improvements in several mental health and well-being measures in the SKY group suggest the effectiveness of the intervention. Moreover, significant improvements in social connectedness were observed, suggesting that SKY could help address the loneliness epidemic in the US. Further, blood sample collection proved to be feasible while simultaneously gathering real-world physiological data, thus enabling exploration of systemic changes in further analysis. These results suggest that the delivery of the SKY intervention is feasible, acceptable, and potentially effective in improving mental health, well-being, and physiological outcomes. Considering the observed effectiveness of the SKY intervention across several clinically relevant outcomes within this relatively homogeneous population, we propose that future well-powered confirmatory studies that would include broader range of populations to enable the molecular level investigation of these benefits.

## AUTHOR CONTRIBUTIONS

RA and PS established the study protocol. RA, VG, and TM conceived the aims of this pilot feasibility study. VG led the study management and data collection. SK and PS led participant recruitment, coordinated with participants for study visits and intervention delivery, and helped with data management. VG and LF performed descriptive analyses of the feasibility metrics. LF performed the analysis of physiological data. HQ performed the analysis of behavioral survey data. VG wrote the first draft of the manuscript and was mentored by RA and TM. NB and TM guided the overall analysis strategy. SB and KM assisted with data analysis and contributed to writing. All authors reviewed and edited the manuscript. RA led the study team, provided oversight on manuscript preparation, and coordination of the research team. TM provided oversight on statistical analyses, and revised the manuscript. All authors contributed to the interpretation of results, reviewed the manuscript, and agreed with the final version.

## Supporting information

Supplemental Methods and Results

## Data Availability

De-identified individual participant data and statistical/analytic codes will be shared for secondary analyses upon reasonable request to the corresponding authors.

## ACKNOWLEDGEMENTS

We thank the Art of Living Foundation for providing course instructors and helping with marketing for recruitment through their websites and social media platforms. We thank Dr. Suprateek Kundu, MD Anderson, for his help with writing the protocol. We thank Ms. Alekhya Puppala and Ms. Anjani Allada for their assistance with cleanup of recruitment data, and Christos Evangelou, PhD, for providing editorial assistance for this manuscript.

