## Supplemental Methods and Results for "Effects of Sudarshan Kriya Yoga (SKY) Breath Meditation on Heart Rate Variability, Sleep and Mental Health in Healthy Individuals : A Pilot and Feasibility Randomized Controlled Trial"

**Eligibility Screening**

Participants were included if they met all of the following criteria:

1. **Age**: Healthy volunteers over 21 years of age.
2. **Language proficiency**: Minimum 5^th^-grade equivalent competency in English.
3. **Commitment**: Willingness to meet the standards of the study, including agreeing to complete at least 75% of the SKY breath practice follow-up sessions during the 8-week intervention.

Participants were excluded if they met any of the following criteria:

1. **Chronic comorbidities**: Diagnosis of any major medical condition including dementia, severe hypertension, cancer, diabetes, rheumatoid arthritis, asthma or other respiratory illnesses, history of seizure disorder, Crohn's disease, lupus, colitis, dermatitis, fibromyalgia, HIV infection, hepatitis B infection, and hepatitis C infection.
2. **Psychiatric conditions**: Diagnosis of any psychiatric condition, including severe clinical depression, bipolar disorder, schizoaffective disorder, suicidal ideation, and attention-deficit/hyperactivity disorder (ADHD).
3. **Recent psychotic episodes**: History of a major psychotic episode within the last 12 months.
4. **Substance dependence or abuse**: Recent history (within the last 3 months) of consistent substance dependence or abuse, based on self-report.
5. **Recent major surgery**: Major surgery within the last 3 months.
6. **Hormone replacement therapy**: Current use of hormone replacement therapy.
7. **Lithium medication**: Current use of lithium or lithium-based medications.
8. **Pregnancy or breastfeeding**: Currently pregnant, breastfeeding, or actively trying to become pregnant.
9. **Body mass index (BMI)**: BMI greater than 35.
10. **Smoking or substance use**: Current smoking (including cigarettes, vapes, hookah) or heavy alcohol or substance use, defined by the CDC as 8+ or 15+ drinks per week for women and men, respectively.
11. **Concurrent study participation**: Concurrent participation in another pilot study or clinical trial of a mind or body intervention.
12. **Existing meditation or breath-based practice**: Existing regular practice (3 or more times a week) of a formal meditation or breath-based technique.

**Data Collection**

All participants were provided with a personal Garmin Vivosmart 5 watch and were required to install 2 smartphone applications: Labfront and Garmin Connect. The intervention group participants were prompted to log their daily SKY practice data on the Labfront companion application to monitor their adherence. Participants were requested to open their mobile applications daily to upload the data to secure Garmin and Labfront cloud servers. To overcome technical glitches affecting the log's accuracy, the study coordinator manually verified study adherence at the end of each week via phone calls or texts with participants.

Behavioral surveys were administered via REDCap at baseline (one week before the study began) and 8 weeks post-intervention. Demographic information such as age, gender, race, ethnicity, body weight, and BMI were collected as part of this survey.

Physiological data were obtained from Garmin watches in a manner similar to that for adherence logs. Labfront software was used to process, aggregate, and organize the physiological data.

Blood samples were collected from all participants at baseline (1 week before study initiation), after the 3-day workshop, and after the 8-week study period by phlebotomists in appropriate vacutainers to enable fractionation of serum, plasma, whole blood DNA, whole blood RNA, and peripheral blood mononuclear cells (PBMCs).

As monetary incentives, a total of $400 in three installments was provided to SKY participants, and a total of $150 in two installments was provided to control participants. The incentives were contingent on minimum study adherence for SKY participants (ie, daily home SKY practice 4 times per week and completing at least 6 of 8 weekly long SKY practices during followup) and compliance with data collection procedures for both groups.

**Study Metrics**

*Feasibility Metrics*

Feasibility metrics were evaluated based on the study process and outcomes. The study process was evaluated by analyzing recruitment and refusal rates among those screened for eligibility, as well as the rates of attrition, retention, compliance, and adherence of participants who completed the 8-week program. With respect to wearable watch compliance, participants were instructed to wear the watch as much as possible during the 8 weeks of the study without interfering with their daily activities. Wearing the provided Garmin watch was not required to receive compensation. Wearable watch compliance was assessed daily for both waking and sleeping periods. Participants were considered to wear the device for that day if they had 1) step count data for daily wear compliance or 2) sleep data for wearable compliance during sleep. Adherence to daily SKY practice was defined as performing at least 3 or more sessions weekly during the follow-up period.^1^ For weekly long Kriya, adherence was defined as performing 6 or more sessions (combined in-person and virtual guided group practice) per week.

*Behavioral Outcomes*

Self-reported behavioral outcome measures under 3 broad categories of mental health, psychological thriving, and personality were measured at baseline and post-intervention (week 8 of follow-up) using various questionnaires administered as an electronic survey packet via REDCap. We selected survey instruments that were used in a previous study that evaluated the effects of SKY practice on mental health and psychological thriving.^2^ A full description of the questionnaires and their references are provided in eTable 1. Mental health was evaluated using measures of burnout, stress, negative affect, and symptoms of distress, anxiety, and depression, with lower scores indicating better mental health. Psychological thriving was assessed using measures of psychological well-being, life satisfaction, positive affect, gratitude, self-compassion, mindfulness, optimism, self-esteem, and social connectedness, with higher scores indicating greater thriving. Additionally, coping styles, personality factors, emotional intelligence, and beliefs and opinions about emotions were assessed.

**eTable 1. Measurement Scales for Self-reported Behavioral Outcomes**

| **Outcome measure** | **Survey instrument** | **Response scales** | **Scales and subscales** |
| --- | --- | --- | --- |
| Burnout | Single-Item Measure  of Burnout^3^ | 5-point Likert  1 (I enjoy work and have no symptoms of burnout)  5 (I feel completely burned out and wonder if I can go on) | 1-item score |
| Perceived stress | Perceived Stress Scale  (PSS-10)^4^ | 5-point Likert  1 (Never) to 5 (Very often) | Total score |
| Depression and  anxiety | Mood & Anxiety  Symptom Questionnaire  (MASQ D30)^5^ | 5-point Likert  1 (Not at all) to  5 (Extremely) | *Total score*: Overall  depression and anxiety |
|  |  |  | *Subscales*: General distress, Anxious arousal, Anhedonic depression |
| Psychological  well-being | Psychological Well-Being  (RYFF-18)^6^ | 7-point Likert  1 (Strongly agree) to  7 (Strongly disagree) | *Total score*: Overall well-being |
|  |  |  | *Subscales*: Autonomy, Personal Growth, Environmental Mastery, Positive Relations with Others, Purpose in Life |
| Subjective  well-being | Satisfaction with Life Scale (SWLS)^7^ | 7-point Likert  1 (Strongly agree) to  7 (Strongly disagree) | Total score |
| Positive and  negative emotions | Positive and Negative  Affect Schedule (PANAS)^8^ | 5-point Likert  1 (Very slightly/not at all) to  5 (Extremely) | *Subscales*: Positive Affect,  Negative Affect |
| Gratitude | The Gratitude Questionnaire  Six-Item Form^9^ | 7-point Likert  1 (Strongly agree) to  7 (Strongly disagree) | Total score |
| Self-compassion | Self-Compassion Scale  (SCS-SF)^10^ | 5-point Likert  1 (Almost always) to 5 (Almost never) | *Total score*:  Overall self-compassion |
| Mindfulness | Five Facets of Mindfulness Questionnaire (FFMQ-15)^11^ | 5-point Likert  1 (Never or very rarely true) to  5 (Very often or always true) | *Total score*: Overall mindfulness |
| Coping styles | Brief-COPE^12^ | 4-point Likert  1 (I haven’t been doing this at all) to  4 (I have been doing this a lot) | *Subscales*: Problem-Focused Coping, Emotion-Focused Coping, Avoidant Coping |
| Optimism | Life Orientation Test  Revised (LOT-R)^13^ | 5-point Likert  1 (Strongly disagree) to  5 (Strongly agree) | Total score |
| Self-esteem | Single-Item Self-Esteem  Scale (SISE)^14^ | 5-point Likert  1 (Not very true of me) to  5 (Very true of me) | 1-item score |
| Social  connectedness | Social Connectedness Scale  Revised (SCS-R)^15^ | 5-point Likert  1 (Strongly disagree) to  5 (Strongly agree) | Total score |
| Eudaimonic  well-being | Subjective Happiness  Scale (SHS)^16^ | 7-point Likert  1 (Not a very happy person/less happy) 7 (A very happy person/more happy) | Total score |
| Personality  factors | Big Five Inventory  (BFI-44)^17,18^ | 5-point Likert  1 (Strongly disagree) to  5 (Strongly agree) | *Domains*: Extraversion, Openness, Agreeableness, Neuroticism, Conscientiousness |
| Emotion mindset | Emotion Mindset Scale^2^ | 6-point Likert  1 (Strongly disagree/never matter)  6 (Completely agree/Always matter) | *Subscales*: Emotional Intelligence, Beliefs/Opinions about Emotion |

*Physiological Measures*

Daily resting heart rate, respiration rate, HRV, and overall sleep scores were obtained over the 8-week study period from data recorded using personal Garmin Vivosmart 5 watches. Daily values for resting heart rate and overall sleep score were precalculated as part of the daily and sleep summary metrics, respectively. Daily values for overall and sleep respiration were determined as the average of all measurements recorded within 24 h and during the sleep period each day, respectively. The root-mean-square of successive differences between normal heartbeats (RMSSD) was used to assess the HRV. Daily values for awake HRV were calculated as the average of all HRV measures recorded during the waking period within a single day, whereas daily values for resting HRV were determined as the average of all measures recorded during the deep-sleep stage within 24 hours. Each participant's waking and deep-sleep periods were determined from their daily records of sleep period data.

**Statistical Analysis**

In the analyses of self-reported behavioral and physiological (Garmin) outcomes, generalized linear models were used to obtain estimates for between-group differences in post-intervention (week 8) measures. All models were adjusted for baseline values. As this was a pilot study, a false discovery rate (FDR) of *P* = .10 was considered appropriate when evaluating the significance of self-reported behavioral outcomes. *P*-values were adjusted for multiple comparisons using the Benjamini and Hochberg (1995) method.^19^ Point estimates for mean differences were reported as the difference in least-square means from the control group (reference), along with 95%, 90%, and 80% confidence intervals for the estimate. In all models for study outcomes, outliers (studentized residual > 3.0 and high leverage on the predictor of interest) were excluded from the analysis. Given the nature of the pilot study, small sample size, and oversampling of intervention participants, we emphasize effect sizes rather than *P*-values to evaluate preliminary effectiveness, although both are reported. Effect sizes were estimated using Hedge’s *g*, which corrects for bias attributed to small sample size and is calculated using the pooled standard deviation.^20^ Estimates for *g* were derived with measures adjusted for baseline values (partial residuals), and values are interpreted as follows: very small (*g* <0.2), small (0.2≤ *g* <0.5), medium (0.5 ≤ *g* <0.8), and large (*g* ≥0.8).

**Supplemental results**

**eTable 2. Adherence to Daily SKY Practice Sessions During Each Week of the Follow-up Period in the SKY Intervention Group (N = 29)**

|  | **Descriptive summaries** | | | | **Participant**  **categories** | | **Adherence**  **status**^a^ | |
| --- | --- | --- | --- | --- | --- | --- | --- | --- |
|  | ***N*** | **Mean (SD)** | **Median**  **(min, max)** | **IQR**  **(Q3 – Q1)** | **≥5**  **sessions** | **≤2**  **sessions** | **High**  **(≥3 sessions)** | **High**  **(≥4 sessions)** |
| **Week 1**  (6/26 – 7/03) | 29 | 4.55 (1.06) | 4.0  (3.0, 6.0) | 1.0  (5.0 – 4.0) | 14 (48.3%) | 0 | 29  (100%) | 24 (82.8%) |
| **Week 2**  (7/03 – 7/09) | 29 | 4.28 (1.00) | 4.0  (3.0, 6.0) | 1.0  (5.0 – 4.02) | 11 (37.9%) | 0 | 29  (100%) | 22 (75.9%) |
| **Week 3**  (7/10 – 7/16) | 29 | 4.34 (1.14) | 4.0  (2.0, 6.0) | 1.0  (5.0 – 4.0) | 12 (41.4%) | 1 (3.45%) | 28  (96.6%) | 22 (75.9%) |
| **Week 4**  (7/17 – 7/23) | 29 | 4.10 (1.18) | 4.0  (2.0, 6.0) | 2.0  (5.0 – 3.0) | 5 (17.2%) | 1 (3.45%) | 28  (96.6%) | 18 (62.1%) |
| **Week 5**  (7/24 – 7/30) | 29 | 4.14 (1.06) | 4.0  (2.0, 6.0) | 1.0  (5.0 – 4.0) | 10 (34.5%) | 2 (6.90%) | 27  (93.1%) | 22 (75.9%) |
| **Week 6**  (7/31 – 8/06) | 29 | 4.34 (1.14) | 4.0  (2.0, 6.0) | 1.0  (5.0 – 4.0) | 13 (44.8%) | 2 (6.90%) | 27  (93.1%) | 23 (79.3%) |
| **Week 7**  (8/07 – 8/13) | 27 | 4.17 (1.36) | 4.0  (1.0, 6.0) | 2.0  (5.0 – 3.0) | 13 (44.8%) | 2 (6.90%) | 27  (93.1%) | 20 (70.0%) |
| **Week 8**  (8/14 – 8/20) | 27 | 4.21 (1.63) | 4.0  (0, 6.0) | 3.0  (6.0 – 3.0) | 13 (44.8%) | 2 (6.90%) | 27  (93.1%) | 20 (70.0%) |

Data are expressed as N participants (%) unless indicated otherwise.

^a^High adherers: participants who performed SKY daily practice either ≥ 3 or ≥ 4 times weekly (minimum requirements for each category). Low adherers: participants who performed daily practice sessions < 3 or < 4 times weekly.

IQR, interquartile range; Q1, quartile 1; Q3, quartile 3.

**eTable 3. Marginal Estimates From Analyses of Behavioral Outcomes at Week 8 of Follow-up**

| **Outcomes** | **SKY** | | **Control** | |
| --- | --- | --- | --- | --- |
|  | **Adjusted**  **mean** | **95% CI** | **Adjusted**  **mean** | **95% CI** |
| Burnout | 1.82 | 1.58, 2.06 | 2.15 | 1.78, 2.51 |
| Perceived Stress | 20.02 | 18.49, 21.54 | 23.55 | 21.25, 25.84 |
| **Mood and anxiety symptoms (MASQ D30)** |  |  |  |  |
| Overall score | 51.62 | 48.19, 55.05 | 62.10 | 56.95, 67.24 |
| General distress | 24.03 | 22.11, 25.95 | 27.27 | 24.38, 30.15 |
| Anxious arousal | 12.26 | 11.25, 13.26 | 15.76 | 14.24, 17.28 |
| Anhedonic depression | 24.99 | 22.91, 27.08 | 28.10 | 24.97, 31.24 |
| **Positive and negative emotions (PANAS)** |  |  |  |  |
| Positive affect | 37.45 | 35.67, 39.23 | 33.50 | 30.83, 36.17 |
| Negative affect | 15.28 | 13.44, 17.13 | 17.28 | 14.51, 20.05 |
| **Psychological well-being (RYFF-18)** |  |  |  |  |
| Overall score | 107.2 | 104.1, 110.4 | 105.1 | 100.3, 109.8 |
| Autonomy | 17.03 | 16.07, 17.99 | 16.52 | 15.08, 17.97 |
| Environmental mastery | 17.01 | 15.99, 18.03 | 15.74 | 14.21, 17.27 |
| Personal growth | 19.76 | 19.15, 20.37 | 19.71 | 18.80, 20.63 |
| Positive relations with others | 18.25 | 17.48, 19.03 | 18.27 | 17.11, 19.43 |
| Purpose in life | 17.01 | 16.16, 17.86 | 16.89 | 15.61, 18.17 |
| Self-acceptance | 18.16 | 17.32, 19.0 | 17.97 | 16.70, 19.23 |
| Eudaimonic well-being | 11.45 | 10.95, 11.96 | 10.31 | 9.54, 11.08 |
| Life satisfaction | 28.22 | 27.19, 29.25 | 26.84 | 25.28, 28.39 |
| Gratitude | 35.11 | 34.17, 36.06 | 35.41 | 33.98, 36.85 |
| Optimism | 24.25 | 23.13, 25.37 | 22.19 | 20.51, 23.87 |
| Self-esteem | 3.85 | 3.52, 4.18 | 3.41 | 2.91, 3.92 |
| Self-compassion | 45.03 | 42.17, 47.89 | 39.02 | 34.65, 43.40 |
| Social connectedness | 84.53 | 81.57, 87.48 | 77.06 | 72.62, 81.50 |
| Mindfulness | 56.31 | 54.59, 58.02 | 52.39 | 49.80, 54.98 |
| **Coping styles (Brief-COPE)** |  |  |  |  |
| Avoidant coping | 10.60 | 9.94, 11.25 | 11.41 | 10.43, 12.39 |
| Problem-focused | 23.78 | 22.69, 24.87 | 22.25 | 20.61, 23.89 |
| Emotion-focused | 27.36 | 26.0, 28.72 | 26.60 | 24.54, 28.65 |
| **Personality factors (BFI-44)** |  |  |  |  |
| Extraversion | 3.41 | 3.31, 3.51 | 3.19 | 3.04, 3.34 |
| Agreeableness | 4.23 | 4.10, 4.37 | 4.07 | 3.87, 4.28 |
| Conscientiousness | 4.04 | 3.90, 4.18 | 3.94 | 3.73, 4.15 |
| Neuroticism | 2.32 | 2.08, 2.56 | 2.69 | 2.33, 3.05 |
| Openness | 3.77 | 3.62, 3.91 | 3.68 | 3.46, 3.90 |
| **Emotion mindset** |  |  |  |  |
| Emotional intelligence | 5.66 | 4.66, 6.66 | 6.35 | 4.85, 7.85 |
| Beliefs/opinions about emotion | 32.19 | 30.46, 33.92 | 32.57 | 29.98, 35.16 |

*^a^*Derived from general linear regression models and adjusted for baseline scores.

BFI-44, Big Five Inventory; Brief-COPE, Coping Orientation to Problems Experienced Inventory; CI, confidence interval; MASQ D30, Mood and Anxiety Symptom Questionnaire; PANAS, Positive and Negative Affect Schedule; RYFF-18, Psychological Well-Being Scale.

**eTable 4. Descriptive Characteristics*^a^* for Physiological Outcomes (Garmin Measures) by Treatment Group at baseline and for Each Day of the SKY Workshop**

|  | **SKY participants** | | | **Control participants** | | |
| --- | --- | --- | --- | --- | --- | --- |
|  | ***N*** | **Mean (SD)** | **Median**  **(min, max)** | ***N*** | **Mean (SD)** | **Median**  **(min, max)** |
| **Resting HR, beats/min** |  |  |  |  |  |  |
| Baseline | 25 | 59.2 (8.25) | 58.5 (43.0, 75.0) | 13 | 60.7 (7.54) | 60.0 (50.0, 76.0) |
| Workshop Day 1 | 25 | 57.4 (6.69) | 58.0 (42.0, 70.0) | 13 | 58.8 (7.27) | 58.0 (49.0, 74.0) |
| Workshop Day 2 | 25 | 57.1 (7.47) | 59.1 (44.0, 70.0) | 13 | 58.6 (5.64) | 56.0 (51.0, 71.0) |
| Workshop Day 3 | 25 | 56.8 (6.98) | 56.0 (45.0, 69.0) | 13 | 60.4 (7.68) | 57.0 (52.0, 78.0) |
| Workshop-Overall | 25 | 57.1 (6.78) | 57.0 (43.7, 67.7) | 13 | 59.3 (6.65) | 57.0 (52.7, 73.0) |
| **Overall RR, breaths/min** |  |  |  |  |  |  |
| Baseline | 25 | 14.5 (0.69) | 14.3 (13.4, 16.0) | 13 | 14.0 (0.48) | 13.9 (13.3, 14.9) |
| Workshop Day 1 | 25 | 14.2 (0.53) | 14.2 (13.2, 15.2) | 13 | 14.1 (0.68) | 14.0 (13.1, 15.4) |
| Workshop Day 2 | 25 | 14.4 (0.74) | 14.4 (13.3, 15.8) | 13 | 14.0 (0.68) | 14.1 (12.8, 15.4) |
| Workshop Day 3 | 25 | 14.5 (0.79) | 14.4 (13.2, 16.2) | 13 | 14.1 (0.71) | 14.1 (13.1, 15.8) |
| Workshop-Overall | 25 | 14.4 (0.63) | 14.4 (13.3, 15.4) | 13 | 14.1 (0.66) | 14.1 (13.0, 15.6) |
| **HRV-Awake** |  |  |  |  |  |  |
| Baseline | 18 | 35.9 (9.32) | 33.8 (21.3, 62.4) | 8 | 32.0 (9.54) | 29.7 (19.2, 48.1) |
| Workshop Day 1 | 16 | 38.4 (10.1) | 36.5 (26.0, 65.7) | 8 | 32.4 (6.92) | 29.3 (25.5, 44.8) |
| Workshop Day 2 | 17 | 35.8 (10.0) | 33.1 (25.4, 60.6) | 6 | 30.0 (8.06) | 25.7 (25.4, 45.6) |
| Workshop Day 3 | 17 | 37.2 (11.0) | 36.2 (24.0, 68.3) | 8 | 30.6 (4.65) | 31.2 (24.1, 37.0) |
| Workshop | 18 | 37.0 (9.83) | 35.9 (25.6, 64.8) | 8 | 31.0 (5.67) | 28.5 (24.8, 40.5) |
| **Sleep RR, breaths/min** |  |  |  |  |  |  |
| Baseline | 20 | 15.3 (2.02) | 14.8 (12.8, 20.5) | 8 | 13.8 (1.12) | 13.8 (12.1, 15.8) |
| Workshop Day 1 | 18 | 15.1 (1.47) | 15.0 (12.7, 18.3) | 8 | 14.1 (1.14) | 13.8 (12.8, 16.3) |
| Workshop Day 2 | 20 | 15.3 (1.80) | 15.1 (13.0, 20.1) | 7 | 14.1 (1.09) | 14.0 (12.2, 15.6) |
| Workshop Day 3 | 20 | 15.2 (1.53) | 14.9 (13.1, 18.1) | 8 | 14.3 (1.14) | 14.3 (12.8, 16.0) |
| Workshop- Overall | 20 | 15.2 (1.50) | 15.2 (12.9, 18.8) | 8 | 14.1 (1.09) | 14.1 (12.6, 16.0) |
| **HRV-sleep** |  |  |  |  |  |  |
| Baseline | 22 | 40.6 (15.4) | 38.0 (18.6, 92.0) | 9 | 35.2 (10.8) | 35.9 (22.1, 60.4) |
| Workshop Day 1 | 19 | 40.8 (14.2) | 41.4 (20.3, 78.9) | 9 | 37.9 (17.2) | 34.9 (15.6, 76.0) |
| Workshop Day 2 | 21 | 41.7 (19.3) | 33.8 (21.1, 88.0) | 8 | 38.0 (11.4) | 36.9 (22.3, 56.3) |
| Workshop Day 3 | 22 | 41.6 (13.6) | 39.7 (22.8, 71.5) | 9 | 31.2 (8.90) | 28.6 (19.2, 45.4) |
| Workshop-Overall | 22 | 41.4 (13.9) | 36.4 (21.4, 68.3) | 9 | 34.9 (12.3) | 32.0 (17.4, 57.1) |
| **Overall sleep score** |  |  |  |  |  |  |
| Baseline | 19 | 72.5 (14.3) | 74.0 (46.0, 94.0) | 8 | 70.6 (13.2) | 71.9 (42.0, 87.0) |
| Workshop Day 1 | 17 | 72.2 (17.8) | 81.0 (38.0, 94.0) | 8 | 75.5 (16.5) | 81.0 (46.0, 90.0) |
| Workshop Day 2 | 19 | 73.4 (19.2) | 82.0 (34.0, 91.0) | 6 | 76.0 (16.1) | 77.5 (48.0, 98.0) |
| Workshop Day 3 | 19 | 70.5 (15.3) | 73.0 (44.0, 93.0) | 8 | 71.0 (20.4) | 78.0 (37.0, 94.0) |
| Workshop-Overall | 19 | 72.2 (12.1) | 76.0 (55.0, 90.7) | 8 | 73.5 (15.3) | 74.7 (44.0, 91.5) |

^a^For comparison purposes, only participants with baseline data were included in the descriptive characteristics.

RHR, resting heart rate; HRV, heart rate variability; RR, respiration rate.

**eTable 5. Descriptive Characteristics*^a^* for Physiological Outcomes (Garmin Measures) for Each Week of Follow-up**

|  | **SKY participants** | | | **Control participants** | | |
| --- | --- | --- | --- | --- | --- | --- |
|  | ***N*** | **Mean (SD)** | **Median (min, max)** | ***N*** | **Mean (SD)** | **Median (min, max)** |
| **Resting HR, beats/min** |  |  |  |  |  |  |
| Week 1 | 25 | 57.4 (7.02) | 58.0 (41.7, 67.6) | 13 | 60.7 (7.10) | 59.0 (52.8, 76.5) |
| Week 2 | 25 | 57.0 (6.47) | 57.0 (42.6, 68.9) | 13 | 59.7 (7.18) | 59.3 (51.2, 78.0) |
| Week 3 | 25 | 56.7 (6.75) | 56.9 (42.9, 68.1) | 12 | 60.0 (9.09) | 58.4 (47.7, 81.6) |
| Week 4 | 25 | 56.7 (7.44) | 56.3 (42.3, 73.3) | 12 | 58.3 (8.51) | 56.2 (47.7, 80.6) |
| Week 5 | 25 | 56.4 (7.21) | 56.3 (39.6, 72.3) | 12 | 57.9 (8.06) | 56.6 (48.3, 79.0) |
| Week 6 | 25 | 57.2 (7.04) | 59.3 (42.4, 75.3) | 12 | 57.4 (6.40) | 56.3 (49.7, 74.6) |
| Week 7 | 23 | 57.8 (8.62) | 59.1 (41.3, 79.8) | 12 | 57.6 (7.47) | 56.5 (50.5, 77.3) |
| Week 8 | 23 | 57.0 (8.61) | 56.9 (41.3, 83.8) | 12 | 57.7 (6.99) | 56.1 (50.3, 75.7) |
| **Overall RR, breaths/min** |  |  |  |  |  |  |
| Week 1 | 25 | 14.6 (0.76) | 14.5 (13.7, 16.4) | 13 | 14.1 (0.56) | 14.1 (13.2, 15.1) |
| Week 2 | 25 | 14.6 (0.73) | 14.6 (13.4, 16.0) | 13 | 14.1 (0.61) | 14.0 (12.9, 15.1) |
| Week 3 | 25 | 14.4 (0.71) | 14.2 (13.4, 15.9) | 12 | 14.1 (0.51) | 14.1 (13.1, 14.8) |
| Week 4 | 25 | 14.6 (0.72) | 14.4 (13.2, 15.9) | 12 | 14.0 (0.56) | 13.9 (12.9, 14.7) |
| Week 5 | 25 | 14.5 (0.66) | 14.4 (13.4, 16.0) | 12 | 14.1 (0.66) | 14.0 (12.6, 14.9) |
| Week 6 | 25 | 14.5 (0.61) | 14.3 (13.5, 16.0) | 12 | 14.0 (0.60) | 14.0 (12.5, 14.8) |
| Week 7 | 23 | 14.6 (0.71) | 14.4 (13.3, 15.9) | 12 | 14.1 (0.71) | 14.1 (12.7, 14.9) |
| Week 8 | 23 | 14.4 (0.74) | 14.4 (13.1, 15.8) | 12 | 14.1 (0.63) | 14.1 (12.9, 15.0) |
| **HRV-awake** |  |  |  |  |  |  |
| Week 1 | 18 | 34.1 (10.8) | 32.8 (22.5, 69.8) | 8 | 29.9 (5.96) | 28.2 (22.5, 40.5) |
| Week 2 | 18 | 35.1 (11.1) | 32.4 (21.7, 69.3) | 8 | 32.2 (8.08) | 29.6 (22.0, 45.3) |
| Week 3 | 17 | 35.7 (11.0) | 34.2 (23.2, 68.9) | 8 | 31.9 (9.16) | 30.7 (20.6, 47.7) |
| Week 4 | 16 | 37.3 (11.9) | 35.6 (24.4, 74.1) | 7 | 33.2 (9.86) | 31.6 (20.4, 50.0) |
| Week 5 | 17 | 37.0 (12.2) | 34.0 (23.9, 75.7) | 7 | 32.2 (8.39) | 32.9 (19.9, 46.7) |
| Week 6 | 17 | 37.3 (9.52) | 36.9 (23.1, 60.8) | 6 | 32.1 (9.29) | 30.0 (21.2, 48.9) |
| Week 7 | 15 | 37.0 (12.5) | 36.5 (23.3, 70.2) | 6 | 32.1 (9.04) | 29.7 (20.5, 47.2) |
| Week 8 | 13 | 39.5 (13.8) | 37.0 (24.6, 77.7) | 6 | 34.5 (7.98) | 34.2 (20.7, 42.6) |
| **Sleep RR, breaths/min** |  |  |  |  |  |  |
| Week 1 | 20 | 15.4 (1.47) | 15.5 (13.4, 17.8) | 8 | 14.1 (0.91) | 14.1 (12.9, 15.4) |
| Week 2 | 20 | 15.5 (1.62) | 15.3 (12.6, 19.1) | 8 | 14.0 (1.10) | 14.0 (12.3, 15.5) |
| Week 3 | 19 | 15.2 (1.55) | 15.0 (12.9, 17.9) | 8 | 14.0 (0.82) | 14.1 (12.7, 15.0) |
| Week 4 | 18 | 15.5 (1.73) | 15.3 (12.9, 18.4) | 8 | 13.8 (0.96) | 13.8 (12.3, 15.1) |
| Week 5 | 19 | 15.5 (1.58) | 15.5 (13.1, 18.6) | 8 | 13.9 (1.13) | 14.1 (11.7, 15.2) |
| Week 6 | 19 | 15.3 (1.35) | 15.4 (13.0, 17.7) | 8 | 13.8 (1.09) | 14.2 (11.5, 15.1) |
| Week 7 | 18 | 15.3 (1.52) | 15.2 (12.8, 18.7) | 8 | 13.8 (1.12) | 14.0 (11.6, 15.1) |
| Week 8 | 17 | 15.3 (1.52) | 15.1 (12.9, 18.6) | 8 | 14.0 (1.10) | 14.4 (12.1, 15.1) |
| **HRV-sleep** |  |  |  |  |  |  |
| Week 1 | 22 | 41.3 (16.6) | 38.1 (21.0, 85.6) | 9 | 34.4 (11.4) | 32.0 (21.6, 58.4) |
| Week 2 | 22 | 43.7 (15.0) | 42.4 (22.8, 76.0) | 9 | 34.7 (13.1) | 33.5 (18.2, 63.5) |
| Week 3 | 21 | 45.5 (20.6) | 39.2 (21.0, 98.0) | 9 | 36.5 (12.6) | 35.3 (17.8, 59.2) |
| Week 4 | 20 | 47.9 (20.6) | 44.5 (23.4, 96.0) | 8 | 36.9 (12.3) | 36.8 (18.0, 53.0) |
| Week 5 | 20 | 46.6 (18.6) | 44.2 (23.7, 90.9) | 8 | 37.3 (13.7) | 38.4 (18.2, 62.7) |
| Week 6 | 20 | 45.0 (17.3) | 40.9 (23.1, 84.6) | 8 | 37.6 (15.7) | 37.7 (19.9, 70.3) |
| Week 7 | 18 | 43.4 (19.4) | 35.8 (19.7, 90.4) | 7 | 33.0 (11.0) | 31.4 (18.5, 46.1) |
| Week 8 | 17 | 47.8 (20.3) | 44.4 (21.3, 98.5) | 7 | 38.7 (13.3) | 36.2 (19.8, 62.2) |
| **Overall sleep score** |  |  |  |  |  |  |
| Week 1 | 19 | 67.4 (12.5) | 67.0 (35.6, 88.9) | 8 | 69.3 (14.7) | 70.8 (41.8, 88.3) |
| Week 2 | 19 | 67.8 (10.9) | 67.4 (47.0, 88.3) | 8 | 67.6 (14.6) | 73.1 (43.0, 80.8) |
| Week 3 | 18 | 64.2 (11.6) | 67.1 (45.9, 82.7) | 8 | 65.1 (14.2) | 62.2 (46.0, 86.3) |
| Week 4 | 17 | 70.2 (9.67) | 70.2 (47.7, 86.6) | 8 | 69.5 (15.5) | 74.5 (43.7, 87.1) |
| Week 5 | 18 | 69.2 (10.2) | 68.9 (50.0, 86.9) | 8 | 66.2 (17.8) | 73.4 (42.4, 85.0) |
| Week 6 | 18 | 67.8 (12.9) | 68.5 (44.0, 96.0) | 8 | 71.3 (11.9) | 71.6 (51.6, 83.6) |
| Week 7 | 17 | 65.2 (10.3) | 65.6 (44.8, 84.4) | 8 | 72.3 (14.2) | 78.4 (42.9, 85.2) |
| Week 8 | 16 | 71.2 (13.2) | 71.1 (46.1, 88.6) | 8 | 65.1 (11.6) | 65.1 (47.4, 80.7) |

^a^For comparison purposes, only participants with baseline data were included in the descriptive characteristics of the workshop metrics.

HR, heart rate; HRV, heart rate variability; RR, respiration rate.

**eTable 6. Sample Sizes and Marginal Estimates From Analyses of Garmin Measures**

|  | **SKY***^a^* | | | **Control***^a^* | | |
| --- | --- | --- | --- | --- | --- | --- |
|  | ***N*** | **Adjusted mean** | **95% CI** | ***N*** | **Adjusted mean** | **95% CI** |
| **Resting HR, beats/min** |  |  |  |  |  |  |
| Workshop Day 3 | 25 | 57.26 | 55.79, 58.72 | 13 | 59.59 | 57.55, 61.63 |
| Week 8 of Follow-up | 22 | 56.34 | 54.73, 57.96 | 12 | 56.76 | 54.57, 58.96 |
| **Overall RR, breaths/min** |  |  |  |  |  |  |
| Workshop Day 3 | 24 | 14.33 | 14.13, 14.53 | 13 | 14.43 | 14.15, 14.72 |
| Week 8 of Follow-up | 22 | 14.29 | 14.11, 14.48 | 12 | 14.32 | 14.07, 14.58 |
| **HRV-awake** |  |  |  |  |  |  |
| Workshop Day 3 | 15 | 33.55 | 31.45, 35.65 | 8 | 31.57 | 28.68, 34.45 |
| Week 8 of Follow-up | 12 | 36.09 | 33.25, 38.94 | 6 | 34.83 | 30.81, 38.85 |
| **Sleep RR, breaths/min** |  |  |  |  |  |  |
| Workshop Day 3 | 19 | 14.98 | 14.67, 15.29 | 8 | 15.01 | 14.53, 15.49 |
| Week 8 of Follow-up | 17 | 14.89 | 14.57, 15.2 | 8 | 14.68 | 14.23, 15.14 |
| **HRV-sleep** |  |  |  |  |  |  |
| Workshop Day 3 | 22 | 40.42 | 37.35, 43.49 | 9 | 33.95 | 29.12, 38.79 |
| Week 8 of Follow-up | 15 | 44.71 | 41.06, 48.36 | 7 | 39.16 | 33.81, 44.5 |
| **Overall sleep score** |  |  |  |  |  |  |
| Workshop Day 3 | 19 | 70.44 | 62.34, 78.53 | 8 | 71.22 | 58.73, 83.7 |
| Week 8 of Follow-up | 23 | 74.43 | 68.38, 80.48 | 12 | 65.55 | 57.5, 73.6 |

CI, confidence interval; RHR, resting heart rate; HRV, heart rate variability; RR, respiration rate

**eTable 7. Wearable Garmin Watch Compliance at the Baseline, Workshop, and Follow-up Time Points^a^**

| **Wearable compliance metrics** | **Overall**  **(N = 42)** | **SKY**  **(N = 29)** | **Control**  **(N = 13)** |
| --- | --- | --- | --- |
| **Device worn ≥ 1 days during the baseline period** |  |  |  |
| While awake (step-count based) | 38 (90.5%) | 25 (86.2%) | 13 (100%) |
| During sleep | 29 (69.0%) | 21 (72.4%) | 8 (61.5%) |
| **Device worn all 3 days of the workshop (SKY participants)** |  |  |  |
| While awake (step-count based) | N/A | 29 (100%) | N/A |
| During sleep | N/A | 21 (72.4%) | N/A |
| **Device worn ≥ 90% of follow-up period (high compliance)** |  |  |  |
| While awake (step-count based) | 39 (92.9%) | 27 (93.1%) | 12 (92.3%) |
| During sleep | 17 (40.5%) | 11 (37.9%) | 6 (46.2%) |
| **Device worn < 70% of follow-up period (low compliance)** |  |  |  |
| While awake (step-count based) | 2 (4.76%) | 1 (3.45%) | 1 (7.69%) |
| During sleep | 12 (28.6%) | 8 (27.6%) | 4 (30.8%) |
| **Total days worn during follow-up***^b^* |  |  |  |
| Mean (SD) | 53.2 (8.07) | 54.5 (3.91) | 50.5 (13.4) |
| Median (min, max) | 55 (6, 56) | 56 (36,56) | 55 (6, 56) |
| Interquartile range [Q3-Q1] | 2 [56-54] | 1 [56-55] | 2 [55-53] |

Data are N participants (%) unless indicated otherwise.

^a^Follow-up period was 56 days for SKY participants and 55 days for control participants.
